# Seroprevalence of SARS-CoV-2 antibodies in an entirely PCR-sampled and quarantined community after a COVID-19 outbreak - the CoNAN study

**DOI:** 10.1101/2020.07.15.20154112

**Authors:** Sebastian Weis, André Scherag, Michael Baier, Michael Kiehntopf, Thomas Kamradt, Steffi Kolanos, Juliane Ankert, Stefan Glöckner, Oliwia Makarewicz, Stefan Hagel, Christina Bahrs, Aurelia Kimmig, Hans Proquitté, Joel Guerra, Bettina Löffler, Mathias W. Pletz, CoNAN study group.

## Abstract

**Background:** Due to the substantial proportion of asymptomatic and mild courses many SARS-CoV-2 infections remain unreported. Therefore, assessment of seroprevalence may detect the real burden of disease. We aimed at determining and characterizing the rate of SARS-CoV-2 infections and the resulting immunity in a defined population.

**Methods:** CoNAN is a population-based cohort study in the previously quarantined community Neustadt-am-Rennsteig, Germany six weeks after a SARS-CoV-2 outbreak with 49 cases identified by PCR screening of all 883 inhabitants. The primary objective of the study was to assess SARS-CoV-2 antibody seroconversion rate using six different IgG detecting immunoassays. Secondary objectives of the study were: *i*.) to determine the rate of seroconversion in children; *ii*.) to determine potential risk factors for symptomatic vs. asymptomatic Covid19 courses; *iii*.) to investigate the rate of virus persistence.

**Findings:** We enrolled 626 participants (71% of the community population). All actual SARS-CoV-2 PCR tests were negative; while a total of 8·4% (52 of 620 tested) had antibodies against SARS-CoV-2 in at least two independent tests. Twenty of the antibody positive participants had previously a positive SARS-CoV-2 PCR. On the contrary, of those 38 participants with SARS-CoV-2 infection, only 20 (52·6%) were antibody positive.

**Interpretation:** Several antibody tests conducted six weeks after an outbreak of SARS-CoV-2 did not detect all previously PCR-positive tested individuals. Cautious evaluation of antibody testing strategies to assess immunity against the infection is warranted.

**Funding:** CoNAN was funded by the Thuringian Ministry for Economic Affairs, Science and Digital Society (TMWWDG).

## INTRODUCTION

SARS-CoV-2 (severe acute respiratory syndrome coronavirus-2) is an emerging pandemic pathogen transmitted by smear, droplet and fomite infection ^1,2^. There are neither vaccines nor specific therapies currently available. The rate of asymptomatic infections is unclear and most of the symptomatic cases take a mild course. However, approximately 15% of the patients and especially older individuals develop a severe disease, *i*.*e*. progressive pneumonia and multi-organ failure that is associated with increased mortality ^1,3^.

Non-medical measures to prevent the spread of SARS-CoV-2 are currently based on the interruption of infection chains through “social distancing", public masking, school closure and reduction of public life (“lockdown”). These have proven to be effective ^4^, yet, they are also associated with substantial social and economic impact. Therefore, a “lockdown”-approach will only be accepted by the societies as long as the advantages, *i*.*e*. protection of those most vulnerable to severe courses of the disease surpass its associated disadvantages ^5^. However, it needs to be taken into account that an early “exit” out of the lockdown is likely to be associated with increasing infection rates that could result in a “second wave of infection”. Hence, the consequences of such a “lockdown exit”, will depend on the extent of the population that remains vulnerable to severe disease courses. It has been argued, that the risk to acquire the infection is minimized if a large percentage of the population has been infected with SARS-CoV-2 and has, at least partially, developed immunity against it ^6^, which is referred to as herd immunity ^5^. Several population-based cohort studies have therefore tried to determine the proportion of infected persons by measuring the sero-prevalence of anti-SARS-CoV2 antibodies. Most of these studies have used only one or two different antibody assays and omitting infants, whom to include is a challenge in such studies. The largest bias of sero-prevalence studies is probably caused by the antibody assays used. Methodology papers have shown that there are tremendous differences between the currently available SARS-CoV-2 antibody assays with a test specificity ranging from 84·3-100% in pre-COVID-19 specimens and inter-test agreements ranged from 75·7-94·8% ^7,8^.

To address some of the constraints, we aimed at determining and characterizing the rate of SARS-CoV-2 infections and the resulting immune responses in a defined population. We chose a population-based approach including infants and used six different IgG antibody assays in parallel. The study was conducted in Neustadt-am-Rennsteig, a village in the Ilm district in central Thuringia, Germany with 883 inhabitants in which a SARS-CoV-2 outbreak had occurred. On March 22^nd^, 11 confirmed Covid-19 cases had been diagnosed in the district of which 6 (55%) were Neustadt residents with further 69 residents classified as contact persons. As a consequence, local public health authorities declared a 14-day quarantine on the entire village in which residents were also not allowed to leave the village. With support of the local family physician, an outbreak containment team of the public health department conducted a mandatory mass screening using nasopharyngeal swabs starting on April 1^st^ in which 865 SARC-CoV-2 PCR tests were performed resulting in the diagnosis of overall 49 SARS-CoV-2 infections. With the initiated containment measures, the outbreak was controlled and the transmission to neighboring villages was prevented. There were three SARS-CoV-2 associated deaths. Due to the isolated location of the village and the clear and controlled outbreak, Neustadt-am-Rennsteig is well suited to study the sero-conversion and immunity of SARS-CoV-2 infections.

## METHODS

### Study design and enrollment

The CoNAN study (Covid-19 outbreak in Neustadt-am-Rennsteig) is an ongoing exploratory population-based cohort study. We here report the baseline characteristics of the participants at the time of the outbreak/quarantine initiation and at study initiation. Follow-up assessments are planned after 6 and 12 months relative to baseline assessment. All households in the community of Neustadt-am-Rennsteig were informed by mail prior to study initiation about the aims of study. Study participation is voluntary and can be withdrawn at any time, refusal to participate has no consequences. Participants were enrolled from May 12^th^ to 16^th^ 2020 at a central study site that was set-up in the villages’ town hall and additional until May 22^nd^ by home visits. After informed consent, questionnaires, blood samples and pharyngeal washes were directly taken at the study site. Pediatricians were part of the study team to adequately involve under-aged participants and to ensure their appropriate sampling as well. At the study site, plasma was directly centrifuged at 4°C/2,000 *g* for 10 minutes and stored at 8°C. Pharyngeal washes were obtained after a short mouth wash with non-sparkling water under direct supervision of a study team member to ensure appropriate quality. Samples were transported twice a day to the Jena University Hospital allowing a timely further processing at the participating research institutes. Participants who could not come to the study site were enrolled by the local primary care physician at their respective homes until the 22^nd^ of May.

### Ethics review, data protection and data management

The study was conducted according to the current version of the Declaration of Helsinki and has been approved by the institutional ethics committees of the Jena University Hospital and the respective data protection commissioner (approval number 2020-1776) and the ethics committee of the Thuringian chamber of physicians. All data were collected with unique pseudonyms on paper case report forms. These identifiers were later used to merge the questionnaire information with the laboratory information in an electronic study database. Study registrations was applied at the German Clinical Trials Register: DRKS00022416.

### Inclusion Criteria

All inhabitants of the community of Neustadt-am-Rennsteig regardless of age, gender or infections status were eligible for participation. Informed consent was provided by the participants or the parents/legal representatives.

### Exclusion Criteria

Individuals that do not reside in Neustadt-am-Rennsteig or that live in the adjacent community of Kahlert were not eligible for inclusion.

### Objectives and outcomes

The primary objective was to determine the SARS-CoV-2 antibody status (sero-conversion rate) of the population of Neustadt-am-Rennsteig with a defined distance to the end of the quarantine period. SARS-CoV-2 antibody status was defined as “positive” if participants had a positive test result in ≥ 2 of the six antibody tests (details below); otherwise participants were classified as “negative”. The secondary objectives of the study were: *i*.) to determine the rate of seroconversion in children; *ii*.) to determine potential risk factors for symptomatic *vs*. asymptomatic Covid19 courses; *iii*.) to investigate the rate of virus persistence (as part of future follow-up assessments).

### Questionnaire

Participants completed a pseudonymized questionnaire directly at the study site. Clusters were reconstructed using the family name, address and information of household members as provided in the questionnaire. After re-assessing the original paper case report forms, obvious errors were corrected, and duplicated entries were deleted. Plausibility checks of demographic data were performed. Symptoms were noted if reported. Strength and duration of symptoms was not weighted in the analysis of this manuscript. Self-reported information on a positive SARS-CoV-2 PCR test at the time point of the outbreak/quarantine initiation was double-checked with the information by the health department of the Ilm-district if the participants gave their permission on the consent form.

### SARS-CoV-2 RT-PCR

Detection of SARS-CoV-2 in pharyngeal wash samples was performed by RT-PCR amplification of SARS-CoV-2 E-gene and S-gene fragments. 200 µL of the pharyngeal washes were first processed for RNA extraction in the InnuPure C16 using the innuPREP virus DNA/RNA kit (both: Analytik Jena, Jena, Germany). Subsequently, the detection of E- and S-gene of SARS-CoV-2 was performed by using the RealStar SARS-CoV-2 RT-PCR kit 1.0 (altona Diagnostics, Hamburg, Germany) on a Rotor-Gene Q real-time PCR cycler (Qiagen, Hilden, Germany). The amplification protocol consisted of a reverse transcription step at 55°C for 20 minutes, a denaturation step at 95°C for 2 minutes and subsequent 45 cycles at 95°C/55°C/72°C for 15/45/15 seconds, respectively. A positive result was defined as amplification of E- and S-gene in a sample with each cycle threshold value (ct) less than 37. Results from apparently inhibited samples with insufficient internal controls (ct > 37) were verified by using a second RT-PCR test. For these samples, RNA was once again extracted from the original pharyngeal wash specimens via QIASymphony using the QIAsymphony DSP Virus/Pathogen MiniKit (Qiagen) according to manufacturer’s instructions. Subsequently, the RT-PCR step was performed on a LightCycler 480 II (F. Hoffmann-La Roche AG, Basel, Switzerland) using the LightMix Modular Sarbecovirus E-gene kit (TIB MOLBIOL, Berlin, Germany). All steps were performed according to the manufacturer’s instructions.

### SARS-CoV-2 antibody testing

Detection of SARS-CoV-2 IgG antibodies was performed with six different quantification methods, of which two were enzyme-linked immunosorbent assays (ELISA) and four were chemiluminescence-based immunoassays (CLIA/CMIA). In addition, a lateral flow assay (combined IgG/IgM), one IgA (ELISA) and two IgM immunoassays (ELISA and CLIA) were performed that in this setting cannot be directly compared to the IgG immunoassays and will therefore not be reported in this manuscript. All tests were carried out according to manufacturers’ instructions. For detailed information on assay characteristics and instruments used see *Supplementary Table 1*. Sensitivities and specificities are shown as provided by the manufacturer. The following assays were used; EDI Novel Coronavirus SARS-CoV-2 IgG ELISA kit (Epitope Diagnostics Inc., San Diego, USA), SARS-CoV-2 IgG ELISA kit (Euroimmun, Lübeck, Germany), SARS-CoV-2 S1/S2 IgG CLIA kit (DiaSorin, Saluggia, Italy), 2019-nCoV IgG kit (Snibe Co., Ltd., Shenzhen, China), SARS-CoV-2 IgG CMIA kit (Abbott, Chicago, USA) and Elecsys Anti-SARS-CoV-2 kit (Roche, Basel Switzerland).

### Statistical Analysis

#### Sample size considerations

The samples size of the CoNAN-cohort is fixed by the number of inhabitants (n=883) of the community of Neustadt-am-Rennsteig. We aimed at including the population as completely as possible. In addition, we consulted the WHO population-based age-stratified sero-epidemiological investigation protocol for SARS-CoV-2 infection ^9^. On the basis of this recommendation, we estimated that a study with 600 samples (*i*.*e*. an inclusion rate of about 70%) should be sufficient to estimate a (true) seroconversion rate <10%/<20% with an expected margin of error of ± 3%/± 4% (defined by the expected width in percent points of the 95% confidence interval for the seroconversion point estimate using “Confidence interval for proportion using normal approximation (n large)” of nQuery 4·0).

### Data analysis

All statistical analyses were performed in the analysis population sometime stratified by age (adults/children and adolescents) and sero-status from the serological tests. Descriptive analyses included the calculation of mean with standard deviation (SD) and medians with minimum and maximum values for continuous variables, and absolute counts (n, with percentages) for categorical variables. Owing to the great data completeness, we performed no data imputations. As inferential statistics, we applied logistic regression models exploring the associations between the participant-reported symptoms, the SARS-CoV-2 PCR-results of the initial mass testing and the binary serostatus outcome. To adjust estimates for cluster effects between participants living in the same household (derived from their address information) we applied generalized estimation equations (GEE) with exchangeable correlation structure and logistic link function. In addition, we adjusted some of the models for sex and age (linear). Results of logistic GEE models are presented as odds ratio (OR) point and interval estimates. Results are presented such that OR>1 indicate increasing odds for a sero-positive finding with increasing exposures. All confidence intervals (CI) were calculated with 95% coverage. CIs are Wald CIs that are not adjusted for multiple comparisons. Similarly, all reported p-values are unadjusted and two-sided. Due to the explorative nature of the study, we avoided “statistical significance testing”. We used the R Language for Statistical Computing (version 4.0.2; R Core Team 2019: R: A Language and Environment for Statistical Computing. R Foundation for Statistical Computing, Vienna, Austria) for all analyses.

## RESULTS

### Participant characteristics

A total of 626 of the 883 inhabitants (71%) participated in the study. Pharyngeal washes were obtained from 617 (98·6 %) participants at the time of the inclusion. All PCR tests were negative. Plasma samples were obtained from a total of 620 (99%) participants who define the analyzed sample cohort. Of those 620 analyzed participants, 58 (9%) were adolescents and children (<18 years of age at inclusion) and 36 (6%) of these were 12 years of age or younger. *Figure 1* shows a flow-chart of the CoNAN study. Characteristics of the participants are given in *Table 1* and *Supplementary Figure 1*. In four participants the results of the initial PCR-testing during the outbreak could not be revealed. None of these had anti-SARS-CoV2 antibodies.

**Figure 1:**
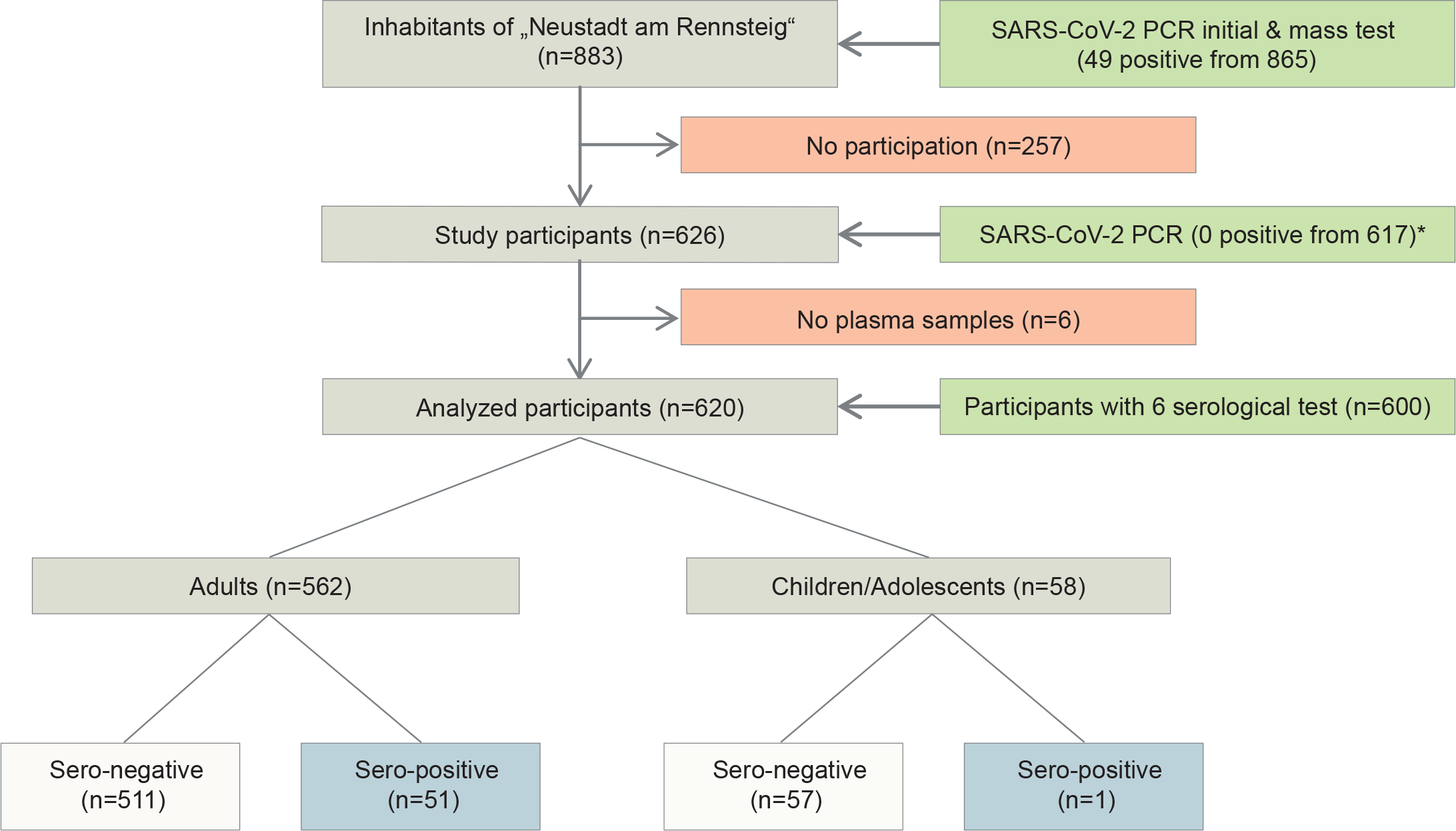
Flow chart of the CoNAN study. * PCR from pharyngeal washes obtained during the CoNAN study in May 2020.

**Table 1:**
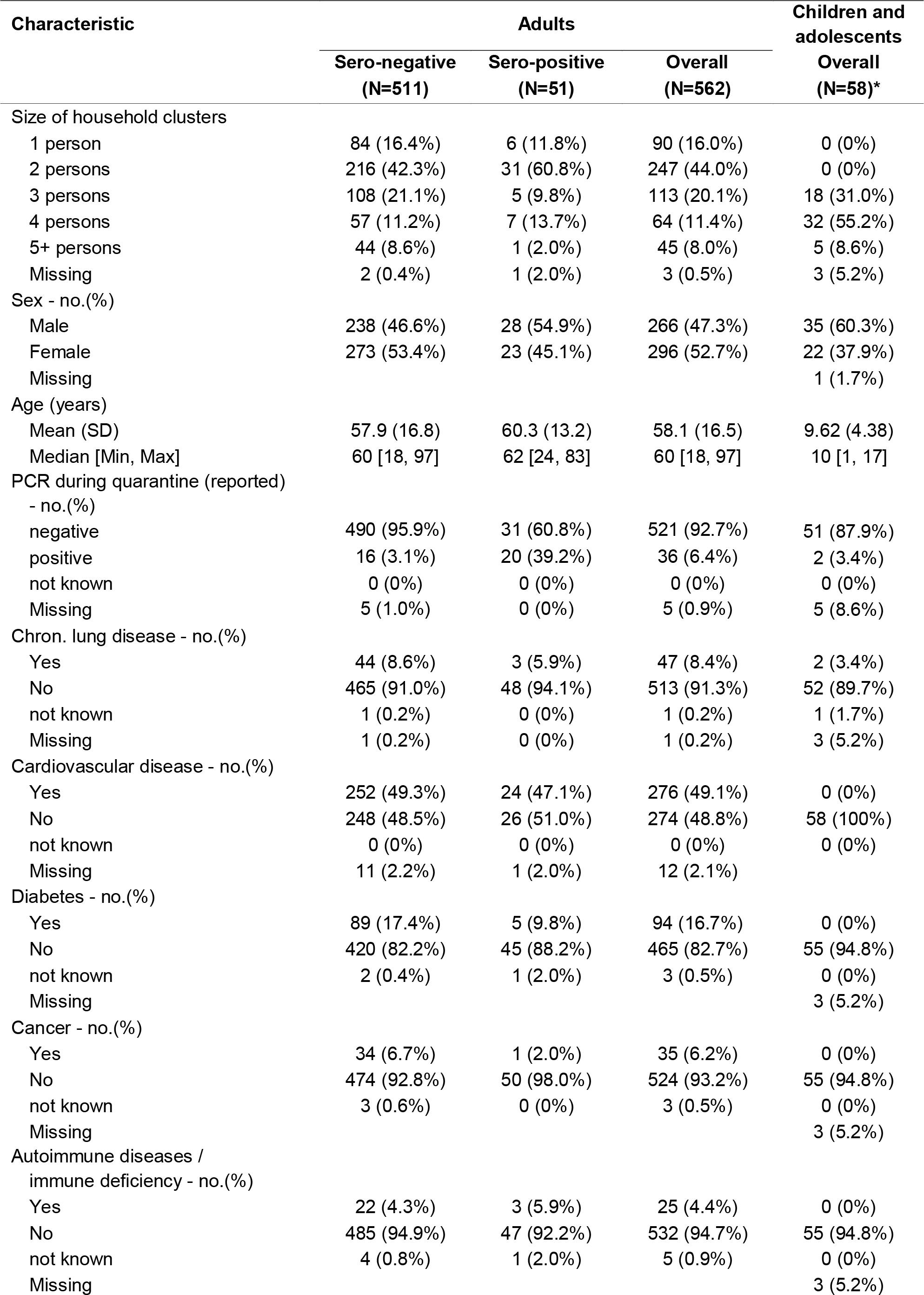
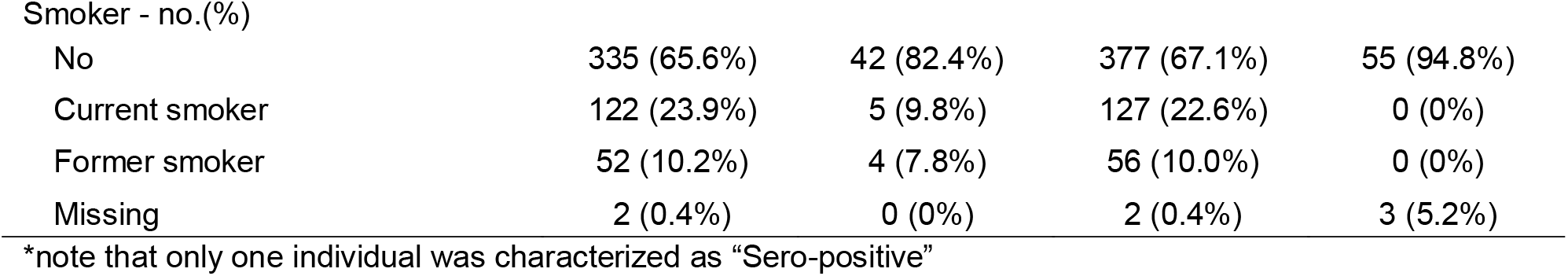
Characteristics of the analyzed (*i*.*e*. with serum samples) 562 adult participants stratified by serostatus and the analyzed (*i*.*e*. with serum samples) 58 participating adolescents and children. Abbreviations: no..number; SD..standard deviation

| Characteristic | Adults |  |  | Children and adolescents |
| --- | --- | --- | --- | --- |
|  | Sero-negative<br>(N=511) | Sero-positive<br>(N=51) | Overall<br>(N=562) | Overall<br>(N=58)* |
| Size of household clusters |  |  |  |  |
| 1 person | 84 (16.4%) | 6 (11.8%) | 90 (16.0%) | 0 (0%) |
| 2 persons | 216 (42.3%) | 31 (60.8%) | 247 (44.0%) | 0 (0%) |
| 3 persons | 108 (21.1%) | 5 (9.8%) | 113 (20.1%) | 18 (31.0%) |
| 4 persons | 57 (11.2%) | 7 (13.7%) | 64 (11.4%) | 32 (55.2%) |
| 5+ persons | 44 (8.6%) | 1 (2.0%) | 45 (8.0%) | 5 (8.6%) |
| Missing | 2 (0.4%) | 1 (2.0%) | 3 (0.5%) | 3 (5.2%) |
| Sex - no.(%) |  |  |  |  |
| Male | 238 (46.6%) | 28 (54.9%) | 266 (47.3%) | 35 (60.3%) |
| Female | 273 (53.4%) | 23 (45.1%) | 296 (52.7%) | 22 (37.9%) |
| Missing |  |  |  | 1 (1.7%) |
| Age (years) |  |  |  |  |
| Mean (SD) | 57.9 (16.8) | 60.3 (13.2) | 58.1 (16.5) | 9.62 (4.38) |
| Median [Min, Max] | 60 [18, 97] | 62 [24, 83] | 60 [18, 97] | 10 [1, 17] |
| PCR during quarantine (reported) |  |  |  |  |
| - no.(%) |  |  |  |  |
| negative | 490 (95.9%) | 31 (60.8%) | 521 (92.7%) | 51 (87.9%) |
| positive | 16 (3.1%) | 20 (39.2%) | 36 (6.4%) | 2 (3.4%) |
| not known | 0 (0%) | 0 (0%) | 0 (0%) | 0 (0%) |
| Missing | 5 (1.0%) | 0 (0%) | 5 (0.9%) | 5 (8.6%) |
| Chron. lung disease - no.(%) |  |  |  |  |
| Yes | 44 (8.6%) | 3 (5.9%) | 47 (8.4%) | 2 (3.4%) |
| No | 465 (91.0%) | 48 (94.1%) | 513 (91.3%) | 52 (89.7%) |
| not known | 1 (0.2%) | 0 (0%) | 1 (0.2%) | 1 (1.7%) |
| Missing | 1 (0.2%) | 0 (0%) | 1 (0.2%) | 3 (5.2%) |
| Cardiovascular disease - no.(%) |  |  |  |  |
| Yes | 252 (49.3%) | 24 (47.1%) | 276 (49.1%) | 0 (0%) |
| No | 248 (48.5%) | 26 (51.0%) | 274 (48.8%) | 58 (100%) |
| not known | 0 (0%) | 0 (0%) | 0 (0%) | 0 (0%) |
| Missing | 11 (2.2%) | 1 (2.0%) | 12 (2.1%) |  |
| Diabetes - no.(%) |  |  |  |  |
| Yes | 89 (17.4%) | 5 (9.8%) | 94 (16.7%) | 0 (0%) |
| No | 420 (82.2%) | 45 (88.2%) | 465 (82.7%) | 55 (94.8%) |
| not known | 2 (0.4%) | 1 (2.0%) | 3 (0.5%) | 0 (0%) |
| Missing |  |  |  | 3 (5.2%) |
| Cancer - no.(%) |  |  |  |  |
| Yes | 34 (6.7%) | 1 (2.0%) | 35 (6.2%) | 0 (0%) |
| No | 474 (92.8%) | 50 (98.0%) | 524 (93.2%) | 55 (94.8%) |
| not known | 3 (0.6%) | 0 (0%) | 3 (0.5%) | 0 (0%) |
| Missing |  |  |  | 3 (5.2%) |
| Autoimmune diseases /<br>immune deficiency - no.(%) |  |  |  |  |
| Yes | 22 (4.3%) | 3 (5.9%) | 25 (4.4%) | 0 (0%) |
| No | 485 (94.9%) | 47 (92.2%) | 532 (94.7%) | 55 (94.8%) |
| not known | 4 (0.8%) | 1 (2.0%) | 5 (0.9%) | 0 (0%) |
| Missing |  |  |  | 3 (5.2%) |
| Smoker - no.(%) |  |  |  |  |
| No | 335 (65.6%) | 42 (82.4%) | 377 (67.1%) | 55 (94.8%) |
| Current smoker | 122 (23.9%) | 5 (9.8%) | 127 (22.6%) | 0 (0%) |
| Former smoker | 52 (10.2%) | 4 (7.8%) | 56 (10.0%) | 0 (0%) |
| Missing | 2 (0.4%) | 0 (0%) | 2 (0.4%) | 3 (5.2%) |

\*note that only one individual was characterized as "Sero-positive"

All six serological tests were performed in 600 (96%) participants. In the remaining 20 individuals (4%), five tests were used for final analysis because, either there was limited material available or the results were inconclusive in one out of the six tests. A comparative performance of the tests is shown in *Figure 2*. In 610 participants, pharyngeal washes and serological test were performed. Upset Plot showing the comparison of test performance between the six serological IgG tests used to evaluate the antibody response in the CoNAN study

**Figure 2:**
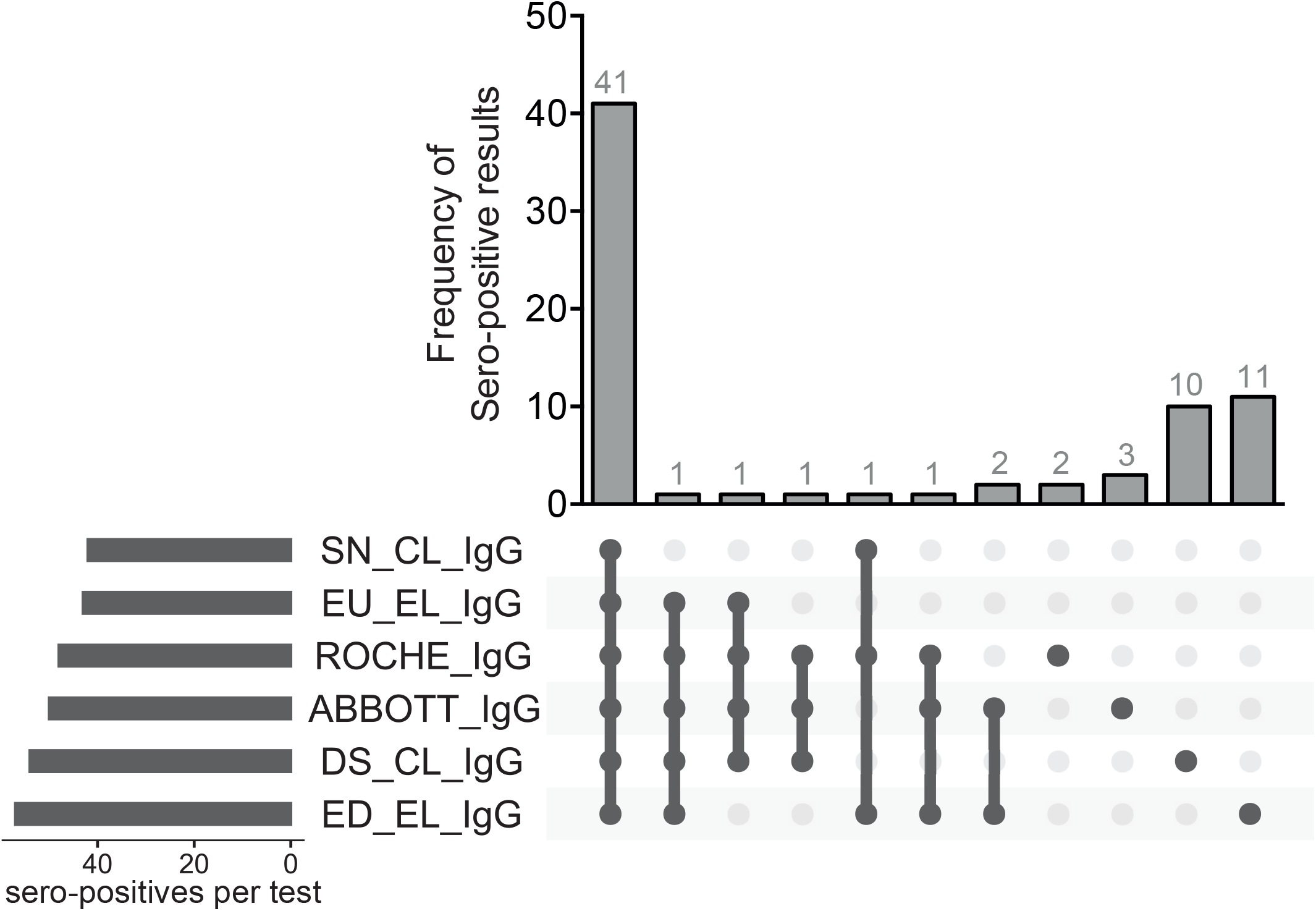
Upset Plot showing the comparison of test performance between the six serological IgG tests used to evaluate the antibody response in the CoNAN study. <u>Abbreviations:</u> DS..SARS-CoV-2 S1/S2 IgG CLIA kit (DiaSorin, Saluggia, Italy); ED..EDI Novel Coronavirus SARS-CoV-2 IgG ELISA kit (Epitope Diagnostics Inc., San Diego, USA); EU..SARS-COV-2 IgG ELISA kit (Euroimmun, Lübeck, Germany); SN.2019-nCoV IgG kit (Snibe Co., Ltd., Shenzhen, China).

### Antibody tests

We found that 52/620 (8·4%) participants had anti-SARS-CoV-2 IgG antibodies of which 20 had been test positive by PCR during the prior sampling at the SARS-CoV-2 outbreak (*Figure 3A; Table 1,2*). Among the antibody-positive participants, there was one child. Therefore, approximately six weeks after proven SARS-CoV-2 infection, antibodies were only detectable in 38·5% participants. Twelve participants with PCR-proven SARS-CoV-2 infection had no symptoms consistent with a respiratory infection or sickness during the last two months, while 180 PCR-negative participants and 168 antibody-negative participants reported respiratory symptoms during the same period, potentially reflecting common respiratory infections in springtime (*Figure 3B*). Thirteen of the 52 seropositive participants (25%) did not report any symptoms of the SARS-CoV-2 infection (*Figure 3C*). Interestingly, two of them; a 55-year old male and a 73-year old male had been tested positive for SARS-CoV-2 infection. In 26 participants, only one out of six serologic tests returned positive. These patients were judged to reflect uncertain cases and assessed as sero-negative for the comparison shown in *Figure 3*. Three of these had previously been tested positive for SARS-CoV-2 infection.

**Table 2:**
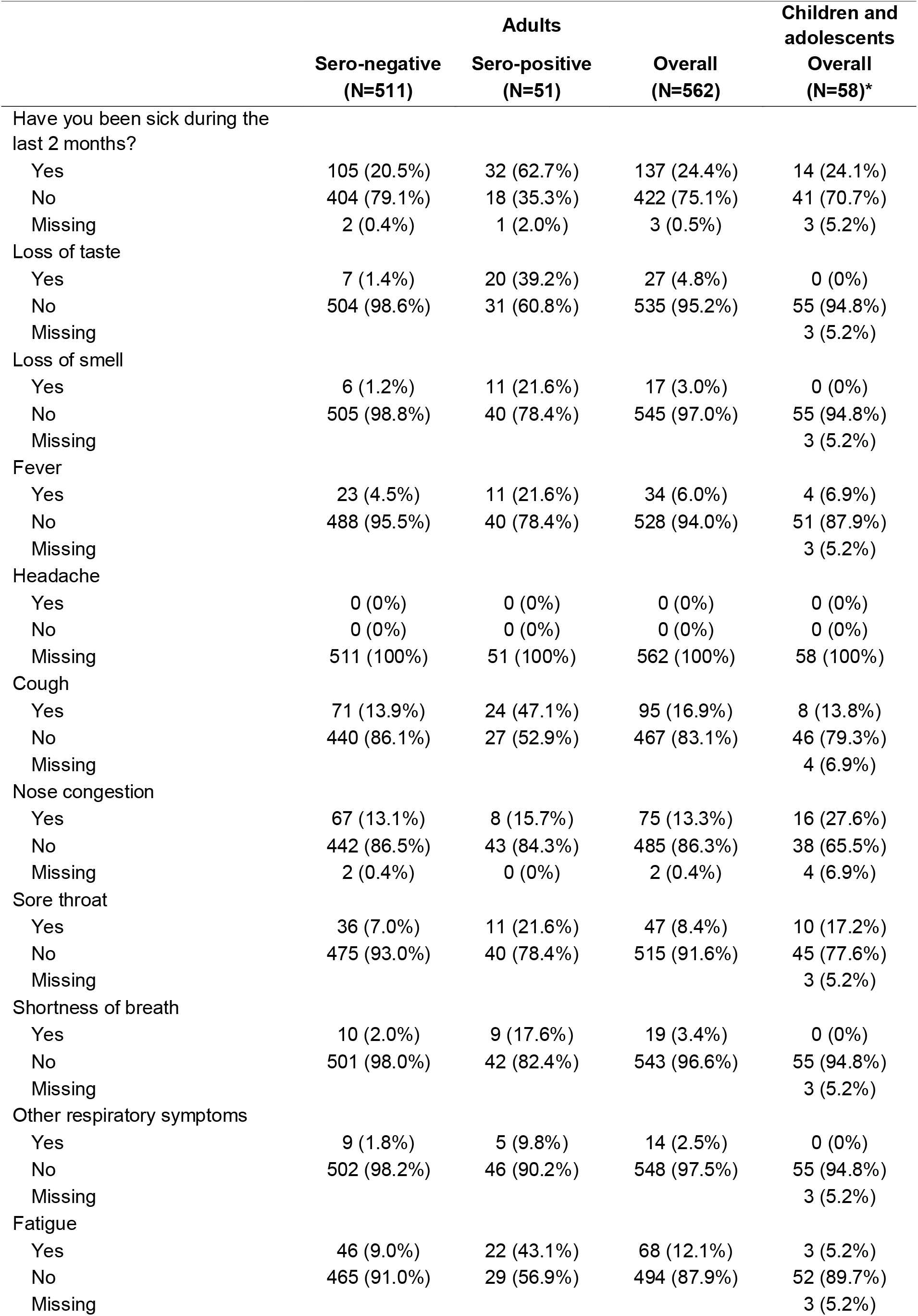
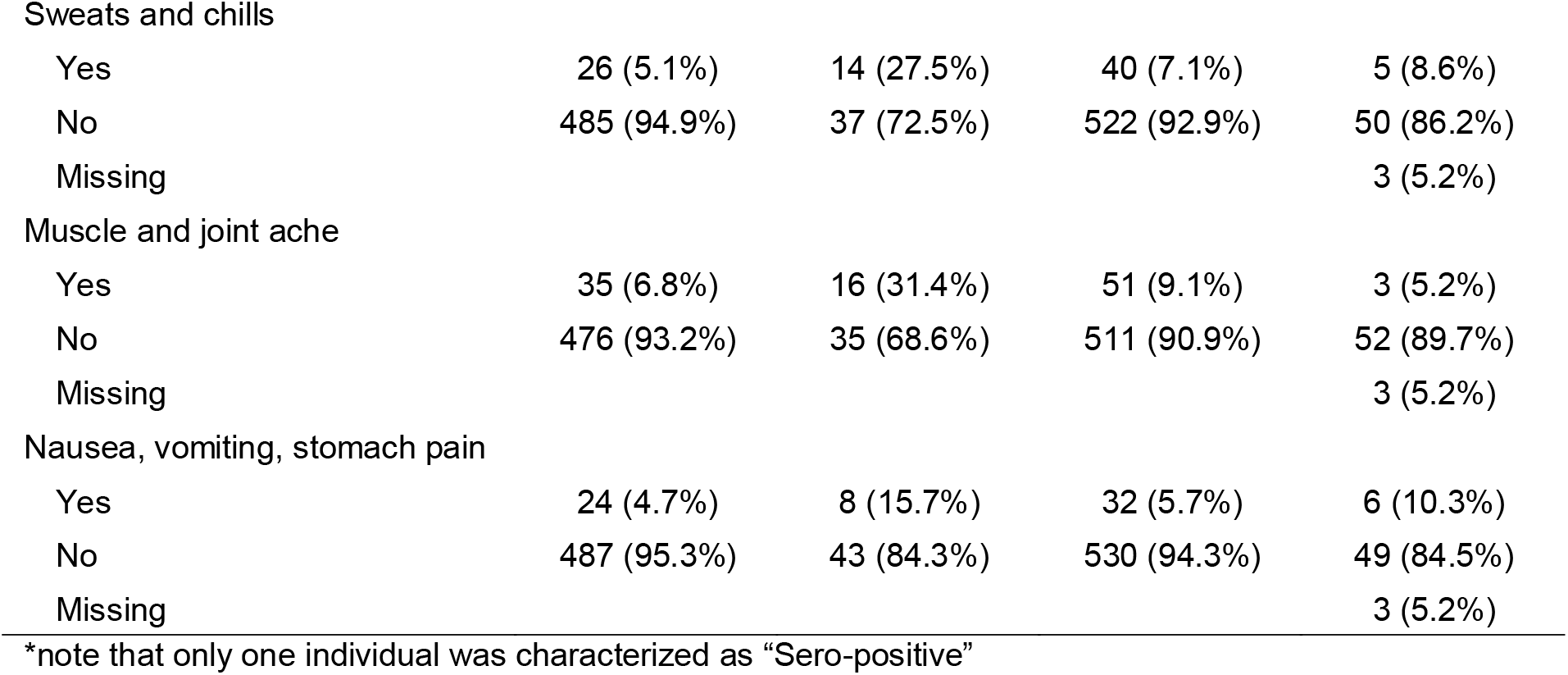
Characteristics of the analyzed (*i*.*e*. with serum samples) 562 adult participants stratified by serostatus and the analyzed (*i*.*e*. with serum samples) 58 participating adolescents and children. Abbreviations: no..number; SD..standard deviation

|  | Sero-negative<br>(N=511) | Adults<br>Sero-positive<br>(N=51) | Overall<br>(N=562) | Children and<br>adolescents<br>Overall<br>(N=58)* |
| --- | --- | --- | --- | --- |
| Have you been sick during the last 2 months? |  |  |  |  |
| Yes | 105 (20.5%) | 32 (62.7%) | 137 (24.4%) | 14 (24.1%) |
| No | 404 (79.1%) | 18 (35.3%) | 422 (75.1%) | 41 (70.7%) |
| Missing | 2 (0.4%) | 1 (2.0%) | 3 (0.5%) | 3 (5.2%) |
| Loss of taste |  |  |  |  |
| Yes | 7 (1.4%) | 20 (39.2%) | 27 (4.8%) | 0 (0%) |
| No | 504 (98.6%) | 31 (60.8%) | 535 (95.2%) | 55 (94.8%) |
| Missing |  |  |  | 3 (5.2%) |
| Loss of smell |  |  |  |  |
| Yes | 6 (1.2%) | 11 (21.6%) | 17 (3.0%) | 0 (0%) |
| No | 505 (98.8%) | 40 (78.4%) | 545 (97.0%) | 55 (94.8%) |
| Missing |  |  |  | 3 (5.2%) |
| Fever |  |  |  |  |
| Yes | 23 (4.5%) | 11 (21.6%) | 34 (6.0%) | 4 (6.9%) |
| No | 488 (95.5%) | 40 (78.4%) | 528 (94.0%) | 51 (87.9%) |
| Missing |  |  |  | 3 (5.2%) |
| Headache |  |  |  |  |
| Yes | 0 (0%) | 0 (0%) | 0 (0%) | 0 (0%) |
| No | 0 (0%) | 0 (0%) | 0 (0%) | 0 (0%) |
| Missing | 511 (100%) | 51 (100%) | 562 (100%) | 58 (100%) |
| Cough |  |  |  |  |
| Yes | 71 (13.9%) | 24 (47.1%) | 95 (16.9%) | 8 (13.8%) |
| No | 440 (86.1%) | 27 (52.9%) | 467 (83.1%) | 46 (79.3%) |
| Missing |  |  |  | 4 (6.9%) |
| Nose congestion |  |  |  |  |
| Yes | 67 (13.1%) | 8 (15.7%) | 75 (13.3%) | 16 (27.6%) |
| No | 442 (86.5%) | 43 (84.3%) | 485 (86.3%) | 38 (65.5%) |
| Missing | 2 (0.4%) | 0 (0%) | 2 (0.4%) | 4 (6.9%) |
| Sore throat |  |  |  |  |
| Yes | 36 (7.0%) | 11 (21.6%) | 47 (8.4%) | 10 (17.2%) |
| No | 475 (93.0%) | 40 (78.4%) | 515 (91.6%) | 45 (77.6%) |
| Missing |  |  |  | 3 (5.2%) |
| Shortness of breath |  |  |  |  |
| Yes | 10 (2.0%) | 9 (17.6%) | 19 (3.4%) | 0 (0%) |
| No | 501 (98.0%) | 42 (82.4%) | 543 (96.6%) | 55 (94.8%) |
| Missing |  |  |  | 3 (5.2%) |
| Other respiratory symptoms |  |  |  |  |
| Yes | 9 (1.8%) | 5 (9.8%) | 14 (2.5%) | 0 (0%) |
| No | 502 (98.2%) | 46 (90.2%) | 548 (97.5%) | 55 (94.8%) |
| Missing |  |  |  | 3 (5.2%) |
| Fatigue |  |  |  |  |
| Yes | 46 (9.0%) | 22 (43.1%) | 68 (12.1%) | 3 (5.2%) |
| No | 465 (91.0%) | 29 (56.9%) | 494 (87.9%) | 52 (89.7%) |
| Missing |  |  |  | 3 (5.2%) |
| Sweats and chills |  |  |  |  |
| Yes | 26 (5.1%) | 14 (27.5%) | 40 (7.1%) | 5 (8.6%) |
| No | 485 (94.9%) | 37 (72.5%) | 522 (92.9%) | 50 (86.2%) |
| Missing |  |  |  | 3 (5.2%) |
| Muscle and joint ache |  |  |  |  |
| Yes | 35 (6.8%) | 16 (31.4%) | 51 (9.1%) | 3 (5.2%) |
| No | 476 (93.2%) | 35 (68.6%) | 511 (90.9%) | 52 (89.7%) |
| Missing |  |  |  | 3 (5.2%) |
| Nausea, vomiting, stomach pain |  |  |  |  |
| Yes | 24 (4.7%) | 8 (15.7%) | 32 (5.7%) | 6 (10.3%) |
| No | 487 (95.3%) | 43 (84.3%) | 530 (94.3%) | 49 (84.5%) |
| Missing |  |  |  | 3 (5.2%) |

\*note that only one individual was characterized as "Sero-positive"

**Figure 3:**
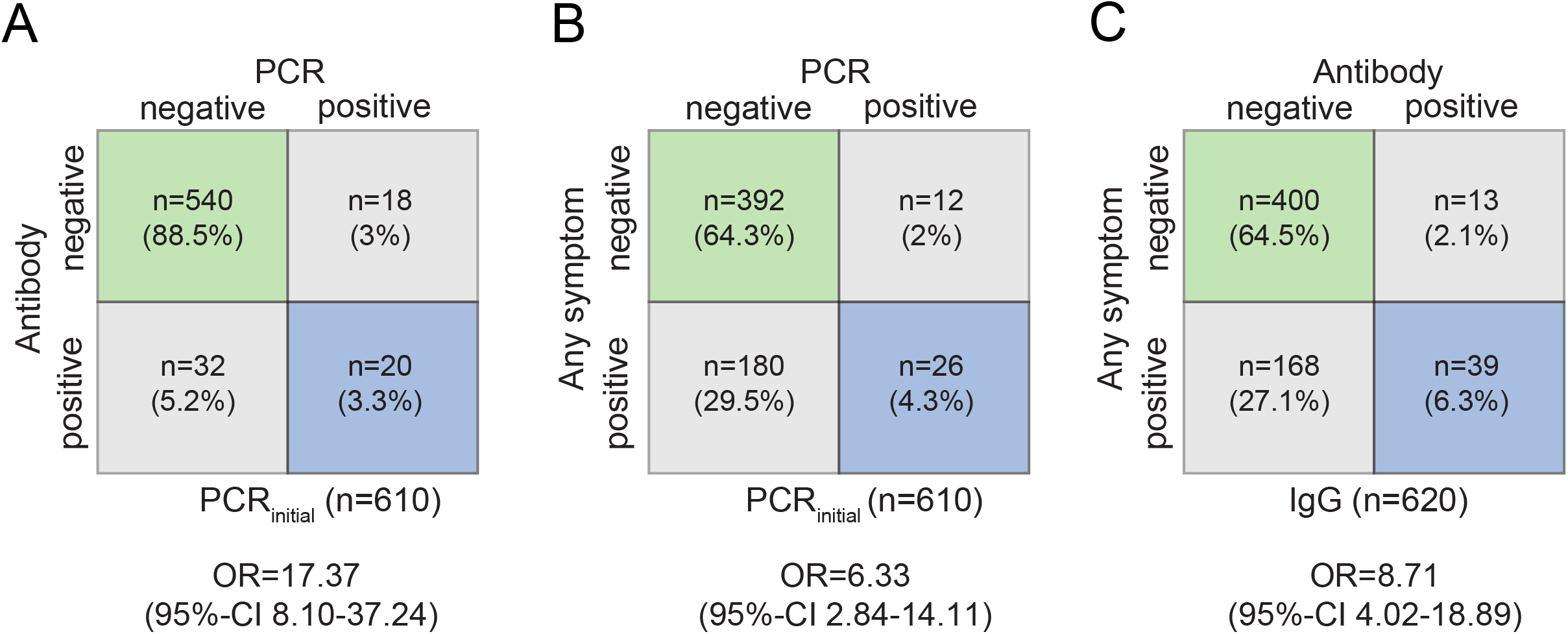
Cross tables of A) antibody status *vs*. SARS-CoV-2 PCR-results (initial mass testing); B) symptoms *vs*. SARS-CoV-2 PCR-results (initial mass testing); C) symptoms *vs*. antibody status. The estimated odds ratios for antibody status (A), any symptoms (B, C) are derived from a logistic GEE model adjusted for sex and age (linear). Note that A) and B) are limited to those 610 participants with an available initial mass-testing PCR-result. Abbreviations: CI..confidence interval, OR..odds ratio.

### Antibody tests and self-reported symptoms

*Figures 3B,C* display a summary of the self-reported symptoms that basically summarizes any of the 14 questions related to symptoms into one variable, *Figure 4* is a more detailed depiction at individual symptoms in all participants (*Figure 4A*) or stratified by the initial SARS-CoV-2 PCR-results (*Figure 4B,C*). Loss of smell and taste were the best predictors of later seropositivity irrespective of stratification with odds ratios point estimates ≥ 10. Interestingly, in individuals that knew they were initially PCR negative, perceived muscle and joint pain, sweats and chills, shortness of breath or fatigue turned out to be predictors of later seropositivity as well. All three investigated variables were strongly associated with OR of 17·37 (95-%CI 8·10-37·24) for PCR *vs*. antibody status, an OR of 6·33 (95%-CI 2·84-14·11) for PCR *vs*. any reported symptom and an OR of 8·71 (95%-CI 4·02-18·89) for antibody status *vs*. any reported symptom, respectively.

**Figure 4:**
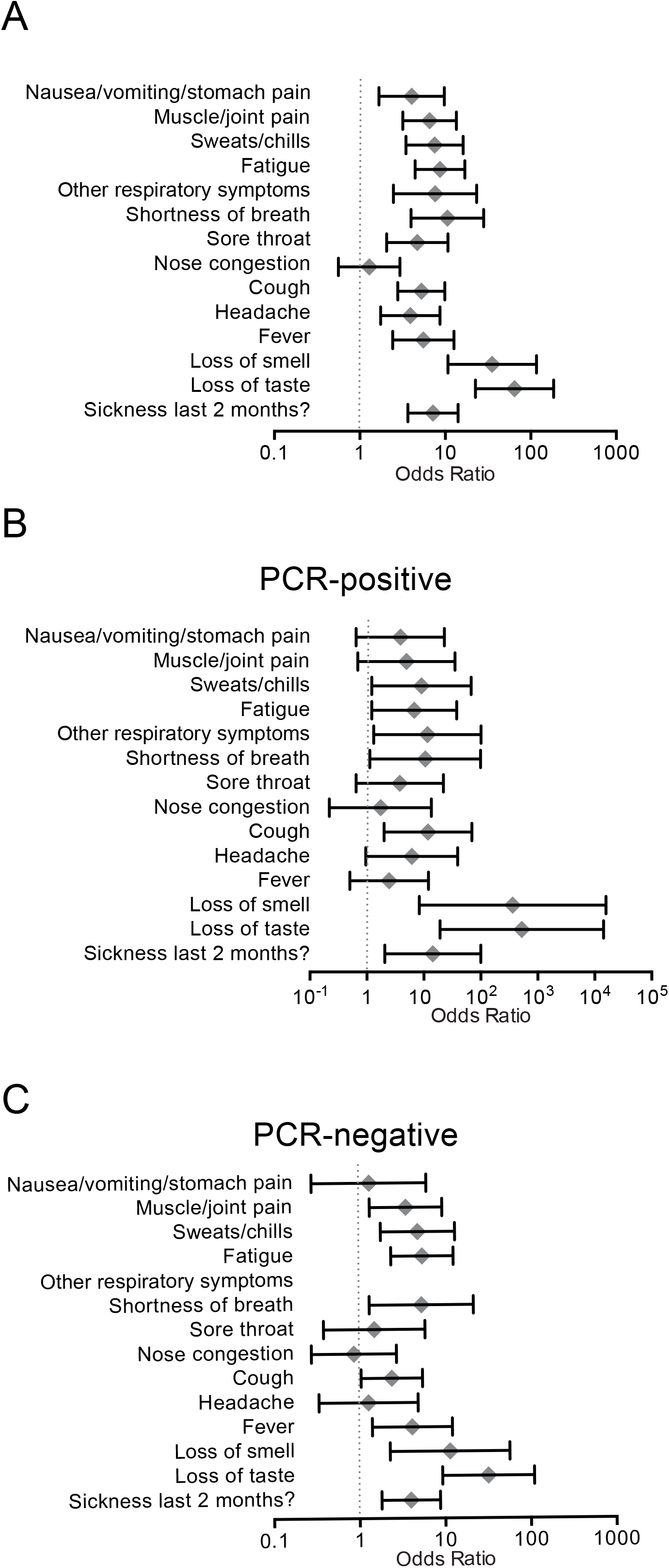
Associations for reported clinical symptoms and positive antibody status for A) all participants, B) previously SARS-CoV-2 PCR-positive and C) previously SARS-CoV-2 PCR-negative. Odds ratio and corresponding 95% confidence interval are derived from the logistic GEE model adjusted for household clustering and sex and age (linear); the plots display the complete cases.

## DISCUSSION

We performed a population-based cohort study enrolling 71% of the population of a central-German village six weeks after a SARS-CoV-2 outbreak with subsequent community quarantining. Our data shows strikingly lower number of seropositive participants than we had expected based on the initial mass screening and the estimates of asymptomatic infections previously reported ^10 11 12^. Only 8·4% of the tested population were seropositive for anti SARS-CoV-2 antibodies in which 6·2% (38/610) had proven SARS-CoV-2 infection, indicating a low rate of asymptomatic cases.

It is currently unknown why in some patients with previously PCR-proven SARS-CoV-2 infection we cannot detect specific antibodies. It has been suggested that less severe clinical manifestations might be associated with lower or absent antibody titers ^13^. However, there are also reports on asymptomatic subjects in whom neutralizing, specific antibodies against SARS-CoV-2 are being found ^14^. Another possibility is that the antibodies were produced, but that the antibody titers declined rapidly, especially as waning of specific antibodies after infection is a common feature observed in corona virus infections ^15,16^. Also, recent data by Long *et al*. suggests that asymptomatic patients might develop weaker immunity against SARS-CoV-2 infection as indicated by an early decrease of IgG and neutralizing antibodies ^17^. Whether the low rate of seroconversion reflects early waning or whether these individuals in fact did not develop antibodies that could be detected with the applied tests, remains to be speculative.

Our post-outbreak seroprevalence cohort studies differs from similar studies ^11,18^ first by the “complete” cohort approach including children and infants instead of a representative sample and second by the extensive use of different antibody assays. An outbreak, similar in median age (58 years) and quarantine measures occurred on the Diamond Princess cruise ship ^18^. In this outbreak, of 3,711 exposed people, there were 619 confirmed SARS-CoV-2 infections corresponding to 17% of which 318 were symptomatic at the time of and 301 had symptoms before testing ^18^(*Table 1*). The infection fatality rate was estimated to be 1·3% (95% CI 0·38-3·6) and the case fatality rate twice higher (2·6%; 95% CI 0·89-6·7) reflecting the 50% of asymptomatic cases. Of note, case- and infection mortality rate dramatically increased in patients of 70 years and older. No data on antibody testing is available for this cohort. Rocklöv *et al*. modeled the effectiveness of infection control measures and suggested that the early intervention prevented 2,000 additional cases ^19^.

Most patients develop antibodies against SARS-CoV-2 within approximately one week after infection ^13^. Several investigators have reported 100% anti-SARS-CoV-2 IgG seropositivity in patients or in covalescent individuals ^20-23^. Using up to six different assays, we found that IgG antibodies were detectable in 39/52 subjects who had had suggestive symptoms of COVID-19 and in 20/38 participants with previously diagnosed SARS-CoV-2 infection. This confirms and extends earlier studies, in which IgG against different SARS-CoV-2 antigens were not detectable in a fraction of patients who were examined at least 14 days after disease onset or convalescents ^11,22,24^. Whereas in some reports the lowest reported rate in convalescent subjects was 77·9% (116/149 subjects) for anti-RBD-IgG and 69·8% (104/149) for anti-S-IgG in a study from New York ^24^.

The significance of the finding that eight participants which reported a transient anosmia or loss of taste that had not previously been tested positive and were antibody negative, remains unclear. Also, the correlation between antibody titers and the level of protection currently is also unknown. Potent neutralizing antibodies have been detected in patients with high or low serum concentrations of antibodies measured by ELISA ^24^ and the level of neutralizing antibodies has been reported to correlate with the number of SARS-CoV-2 specific T-cells ^25^. The correlation between antibodies and protection against COVID-19 is further complicated by evidence suggesting antibody-induced disease enhancement in other coronavirus infections including SARS ^26^. All available evidence indicates that antibody responses alone do not suffice to overcome SARS-CoV-2 infection. Data from SARS-CoV and MERS-CoV suggest that T-cell responses are required for protection and may last longer than antibody titers ^26-30^. Consequently, we are currently analyzing the neutralization capacity in cell culture systems and SARS-CoV-2 specific T-cell responses in our study participants.

### Limitations

Our study has several limitations: *i*.) our study was a population-based cohort study. We were able to recruit 71% of the community population. However, 29% of the population did not participate for unknown reasons which could introduce a bias in the assessment; *ii*.) the study was carried out six weeks after the end of the 14-day quarantine. This could have missed a number of participants that had a rapidly waning antibody response and *iii*.) there was no baseline of the antibody status before the quarantine as some participants might have been exposed earlier during the pandemic.

## CONCLUSIONS

Our data questioned the relevance and reliability of IgG antibody testing to detect past SARS-CoV-2 infections six weeks after an outbreak. We conclude that assessing immunity for SARS-CoV-2 infection should not only rely on antibody tests but might also include the determination of neutralizing antibodies and potentially cellular immunity and requires long term follow up studies.

## Data Availability

The data that support the findings of this study are available from the corresponding author upon reasonable request after publication of the manuscript.

## ABBREVIATIONS

CI: confidence interval
GEE: generalized estimation equations
MERS-CoV: Middle East respiratory syndrome-related coronavirus
OR: odds ratio
SARS-CoV: severe acute respiratory syndrome-related coronavirus

## Acknowledgement

The project is carried out in cooperation with the district administration and the health department of the Ilm-district and is funded by the Thuringian Ministry for Economic Affairs, Science and Digital Society (TMWWDG). We thank Heike Kiesewetter, Anett Büschel and Luzie Rösel for technical assistance.

## REFERENCES

1. Huang C, Wang Y, Li X, et al. Clinical features of patients infected with 2019 novel coronavirus in Wuhan, China. Lancet 2020; 395(10223): 497–506.

2. Whitworth J. COVID-19: a fast evolving pandemic. Trans R Soc Trop Med Hyg 2020; 114(4): 241–8.

3. Chen N, Zhou M, Dong X, et al. Epidemiological and clinical characteristics of 99 cases of 2019 novel coronavirus pneumonia in Wuhan, China: a descriptive study. Lancet 2020; 395(10223): 507–13.

4. Nussbaumer-Streit B, Mayr V, Dobrescu AI, et al. Quarantine alone or in combination with other public health measures to control COVID-19: a rapid review. Cochrane Database Syst Rev 2020; 4: CD013574.

5. Smith CE. Prospects for the control of infectious disease. Proc R Soc Med 1970; 63 (11 Part 2): 1181–90.

6. Randolph HE, Barreiro LB. Herd Immunity: Understanding COVID-19. Immunity 2020; 52(5): 737–41.

7. Whitman JD, Hiatt J, Mowwery CT, et al. Test performance evaluation of SARS-CoV- 2 serological assay. medRxiv preprint 2020; Version 2 (May 17).

8. Jaaskelainen AJ, Kuivanen S, Kekalainen E, et al. Performance of six SARS-CoV-2 immunoassays in comparison with microneutralisation. J Clin Virol 2020; 129: 104512.

9. WHO-2019.https://apps.who.int/iris/bitstream/handle/10665/331656/WHO-2019-nCoV-Seroepidemiology-2020.1-eng.pdf?sequence=1&isAllowed=y. 2019.

10. Hains DS, Schwaderer AL, Carroll AE, et al. Asymptomatic Seroconversion of Immunoglobulins to SARS-CoV-2 in a Pediatric Dialysis Unit. JAMA 2020; 323(23): 2424–5.

11. Streeck H, Schulte B, Kuemmerer B, et al. Infection fatality rate of SARS-CoV-2 infection in a German community with a super-spreading event. medRxiv preprint 2020; Version 2(June 2).

12. Shields AM, Faustini SE, Perez-Toledo M, et al. SARS-CoV-2 seroconversion in health care workers. medRxiv preprint 2020; Version 1(May 19).

13. Huang AT, Garcia-Carreras B, Hitchings MDT, et al. A systematic review of antibody mediated immunity to coronaviruses: antibody kinetics, correlates of protection, and association of antibody responses with severity of disease. medRxiv Preprint 2020; Version 1 (Apr 17).

14. Kraehling V, Kern M, Halwe S, et al. Epidemiological study to detect active SARS- CoV-2 infections and seropositive persons in a selected cohort of employees in the Frankfurt am Main metropolitan area. medRxiv preprint 2020; Version 1(May 25).

15. Callow KA, Parry HF, Sergeant M, Tyrrell DA. The time course of the immune response to experimental coronavirus infection of man. Epidemiol Infect 1990; 105(2): 435–46.

16. Wu LP, Wang NC, Chang YH, et al. Duration of antibody responses after severe acute respiratory syndrome. Emerg Infect Dis 2007; 13(10): 1562–4.

17. Long QX, Tang XJ, Shi QL, et al. Clinical and immunological assessment of asymptomatic SARS-CoV-2 infections. Nat Med 2020.

18. Russell TW, Hellewell J, Jarvis CI, et al. Estimating the infection and case fatality ratio for coronavirus disease (COVID-19) using age-adjusted data from the outbreak on the Diamond Princess cruise ship, February 2020. Euro Surveill 2020; 25(12).

19. Rocklov J, Sjodin H, Wilder-Smith A. COVID-19 outbreak on the Diamond Princess cruise ship: estimating the epidemic potential and effectiveness of public health countermeasures. J Travel Med 2020; 27(3).

20. Sun B, Feng Y, Mo X, et al. Kinetics of SARS-CoV-2 specific IgM and IgG responses in COVID-19 patients. Emerg Microbes Infect 2020; 9(1): 940–8.

21. Jiang HW, Li Y, Zhang HN, et al. Global profiling of SARS-CoV-2 specific IgG/IgM responses of convalescents using a proteome microarray. Nat Commun 2020; 11(1): 3581.

22. To KK, Tsang OT, Leung WS, et al. Temporal profiles of viral load in posterior oropharyngeal saliva samples and serum antibody responses during infection by SARS-CoV-2: an observational cohort study. Lancet Infect Dis 2020; 20(5): 565–74.

23. Xiang F, Wang X, He X, et al. Antibody Detection and Dynamic Characteristics in Patients with COVID-19. Clin Infect Dis 2020; ciaa461.

24. Robbiani DF, Gaebler C, Muecksch F, et al. Convergent antibody responses to SARS-CoV-2 in convalescent individuals. Nature 2020. Jun 18.

25. Ni L, Ye F, Cheng ML, et al. Detection of SARS-CoV-2-Specific Humoral and Cellular Immunity in COVID-19 Convalescent Individuals. Immunity 2020; 52(6): 971–7 e3.

26. Vabret N, Britton GJ, Gruber C, et al. Immunology of COVID-19: Current State of the Science. Immunity 2020; 52(6): 910–41.

27. Liu WJ, Zhao M, Liu K, et al. T-cell immunity of SARS-CoV: Implications for vaccine development against MERS-CoV. Antiviral Res 2017; 137: 82–92.

28. Chen J, Subbarao K. The Immunobiology of SARS*. Annu Rev Immunol 2007; 25: 443–72.

29. Zhao J, Zhao J, Mangalam AK, et al. Airway Memory CD4(+) T Cells Mediate Protective Immunity against Emerging Respiratory Coronaviruses. Immunity 2016; 44(6): 1379–91.

30. Tang F, Quan Y, Xin ZT, et al. Lack of peripheral memory B cell responses in recovered patients with severe acute respiratory syndrome: a six-year follow-up study. J Immunol 2011; 186(12): 7264–8.

